# Neighborhood Socioeconomic Status and Dog-Mediated Rabies: Disparities in Incidence and Surveillance Effort in a Latin American City

**DOI:** 10.1101/2025.04.02.25325110

**Authors:** Sherrie Xie, Julianna Shinnick, Elvis W. Diaz, Edith Zegarra, Ynes Monroy, Sergio E. Recuenco, Ricardo Castillo-Neyra

## Abstract

**Background:** Dog-mediated human rabies is intuitively linked to poverty, but few studies have formally investigated the relationship between local socioeconomic disadvantage and dog rabies incidence.

**Methods:** We leveraged a unique, high-spatial-resolution surveillance database from the rabies-endemic city of Arequipa, Peru to probe the relationship between neighborhood socioeconomic status (SES) and dog rabies risk in 2015-2022. Rabies cases and samples were assigned to the SES level of their block or locality of origin, respectively. We tested the hypothesis that lower SES is associated with increased case positivity, and used a spatial statistical model to understand how sample positivity varied spatially.

**Results:** Rabies cases were concentrated in socioeconomically disadvantaged blocks (*p* < 0·001), and sample positivity had a significant and positive association with neighborhood disadvantage (*p* < 0·05 for all periods), suggesting that surveillance effort was low relative to case incidence in disadvantaged areas. Stratifying samples by those collected via active vs. passive surveillance revealed that active surveillance reduced disparities in surveillance effort and sample positivity. Spatial analysis identified a sample positivity hotspot in a socioeconomically disadvantaged region with low access to health facilities.

**Conclusions:** Dog-mediated rabies is known to impact the poorest regions globally. We found similar patterns mirrored on a much smaller spatial scale - within a single city’s limits. A balanced approach combining spatially-targeted (“active”) and community-based (“passive”) surveillance can help reduce rabies disparities. Mass dog vaccination and surveillance programs could target disadvantaged neighborhoods to decrease inequities in rabies risk to human populations and more effectively control dog rabies epidemics.

## INTRODUCTION

Dog-mediated rabies is a neglected zoonotic disease that causes tens of thousands of human deaths annually.^1,2^ An acute encephalitis caused by a lyssavirus, rabies is 100% fatal once symptoms appear.^3^ It disproportionately affects low and middle-income countries (LMICs), which carry 99% of the global burden of the disease.^2,4^ The vast majority (99%) of human rabies cases are transmitted through dog bites, with most dog-mediated human rabies cases occurring in Southeast Asia and Sub-Saharan Africa.^3,5^ However, dog rabies remains endemic in parts of the Americas, including Haiti, Guatemala, Bolivia, and southern Peru where ongoing transmission of rabies virus in dog populations leads to regular spillover into humans.^6–8^

Dog-mediated human rabies mortality is commonly linked to poverty,^9–11^ partly due to limited access to rabies post-exposure prophylaxis (PEP) by under-resourced communities.^10^ At the cost of USD 65-108 per treatment, PEP has been unaffordable for health systems and communities in many LMICs,^12^ although recent investment from Gavi, the global public-private vaccine alliance, may help bridge this gap.^13^ Even if PEP is available to healthcare systems, it may be inaccessible to people who have experienced dog bites due to lack of availability at nearby health centers,^14,15^ lack of knowledge or misconceptions about PEP as a treatment option,^16^ or distance to health centers.^17^ Additionally, studies conducted in LMICs have found that individuals from low socioeconomic backgrounds are less knowledgeable about rabies and prevention practices, less likely to vaccinate their dogs against rabies, and less likely to seek medical care after a dog bite.^18–26^

In addition to disparities in healthcare access and knowledge of prevention practices, disparities in dog rabies incidence and surveillance may also drive the association between dog-mediated human rabies and poverty. However, studies investigating the relationship between area-level socioeconomic status (SES) and dog rabies incidence have yielded inconsistent results. A spatial analysis of dog rabies in Thailand revealed that more cases occurred in high-poverty areas.^27^ However, a study conducted with data from El Salvador contradicted these findings; the study’s authors concluded that dog rabies risk was higher in the country’s low-poverty zones.^28^ Limitations of both these prior studies include: (1) the use of case data only and a lack of negative samples to provide information about spatial variation in surveillance effort and (2) the use of relatively low-resolution spatial data (i.e., rabies incidence and SES aggregated at the level of districts or municipalities, each comprising an average 20,000-80,000 people).

Here, we leverage a unique high-spatial-resolution surveillance dataset from Arequipa, Peru to understand the relationship between neighborhood SES and dog-mediated rabies risk. Dog rabies has become endemic in Arequipa a few years after the region was declared rabies free, a result of viral reintroduction into the local dog population that was first detected in 2015.^29^ Annual mass dog vaccination campaigns have been held since 2015 to combat the epidemic; however, reaching adequate herd immunity has remained a challenge due to local resource constraints and various barriers to participation faced by dog owners.^30–33^ There is marked socioeconomic inequality in Arequipa, as in many Latin American cities, and we aimed in the present study to determine how the risk posed by dog rabies differentially impacts residents across space and socioeconomic divides. To evaluate the presence of social and spatial inequality in dog rabies burden and surveillance, we test the hypothesis that positivity rates of samples submitted for dog rabies testing increase with low neighborhood SES (i.e., neighborhood disadvantage), map the geospatial distribution of sample positivity, and assess for the presence of positivity hotspots using spatial statistics.

## METHODS

### Data sources

There is a passive surveillance system for dog rabies in Arequipa, Peru, in which dead or suspected dogs are reported by residents to the local Ministry of Health (MoH) either by phone or in-person notification at health facilities (municipal hospitals, health centers, and health posts). Following notification, MoH personnel investigate and collect brain samples from suspected cases for testing by the direct fluorescent antibody test, which is performed using a standardized protocol for rabies diagnosis that is in accordance with WHO guidelines.^34^ Beginning in 2021, the Zoonotic Disease Research Center (a consortium between the University of Pennsylvania and Cayetano Heredia University) has supplemented the passive surveillance system with active surveillance, whereby field personnel search the city’s dry water channels for dead dogs and submit samples from any they find for rabies testing using the protocol described above. Dry water channels are targeted by active surveillance because they are frequently utilized as an ecological corridor by the city’s free-roaming dogs and have been previously associated with the locations of rabid dogs.^29,35^ Precise longitude and latitude coordinates are recorded for all confirmed cases, and the locality (subdistrict division containing 984 people on average) in which a reported dog was found is recorded for samples submitted for testing.

A large geospatial database of all households in Arequipa is maintained by the Zoonotic Disease Research Center that contains for each residential block either: (1) the coordinates of all houses on the block or (2) a block-level aggregate house count. The Peru National Institute of Statistics and Informatics (INEI) determines block-level SES through a national household economic survey that is further imputed using information from the national census using a small-area SES estimation method developed by the World Bank.^36,37^ Block-level SES is calculated based on average estimated household income and further stratified as an ordinal variable using the Dalenius-Hodges method, taking values *A*, *B*, *C*, *D*, *E*, or *NA*.^36,38^ *A* represents the highest (least disadvantaged) SES level and *E* represents the lowest (most disadvantaged) level. Blocks with insufficient data to determine SES are classified *NA*; notably, *NA* assignment often occurs in newly developing, peri-urban areas where households are typically more materially disadvantaged than those in more established areas.^39^ To distinguish the *NA* classification from true missing data, we will refer to this level as *undefined* for the remainder of this manuscript.

### Assigning SES levels to confirmed rabies cases and surveillance samples

Rabies cases were assigned SES levels by intersecting case coordinates with city block polygons and assigning cases to the SES level of the block they fell in. Case coordinates that fell outside of block polygons but within 50 meters of a city block were assigned the SES level of their closest block. Fifty meters was chosen as the distance threshold as it represented the distance within which case coordinates could be clearly associated with a residential area. Case coordinates located more than 50 meters from a city block were not assigned an SES level and were excluded from further analyses. Localities that contained a median of 13 blocks (interquartile range = 7-26), were assigned the median SES level of the blocks within them (after excluding blocks with undefined SES), except for localities in which the majority (>50%) of blocks had undefined SES, which were assigned the *undefined* SES level. All spatial data manipulations were performed in R using the *sf* package.^40–42^ Maps of confirmed case locations, locality-level sample counts, and block-level SES were generated using the *ggmap* R package.^43^

### Evaluating the relationship between sample positivity and neighborhood SES

Sample positivity – which is calculated by dividing the number of confirmed cases for a group or location by the number of samples submitted for that group or location – served as the primary measure by which we evaluated dog rabies disparities, because it combines the burden posed by dog rabies incidence (via confirmed cases) and surveillance effort (via submitted samples). To test the hypothesis that sample positivity increased with neighborhood disadvantage, we performed one-sided Cochran-Armitage trend tests, which can be used to assess for the presence of a trend in the binomial proportions of an outcome variable (sample positivity) across the levels of an ordinal exposure variable (neighborhood SES). To evaluate for consistency of trends over time, we applied the Cochran-Armitage test to samples stratified across two-year intervals: 2015-2016, 2017-2018, etc. Two-year intervals were chosen to capture any broad temporal changes without resulting in overly small cell sizes. We also applied the test across all years to assess for a global trend. We performed the tests by excluding samples assigned to the *undefined* SES level. As a sensitivity analysis, we also conducted the tests by re-assigning *undefined* samples to SES level *E*, as households located in these newly developing areas in the city periphery tend to be the most materially disadvantaged.^39^

We also tested for disparities in the total case count by comparing the counts of cases found in disadvantaged blocks (*i.e.*, SES levels *D* or *E*) and less disadvantaged blocks (*i.e.*, SES levels *A*, *B*, or *C*) to the counts of households within the city that fall within these same SES groups using a chi-squared test with one degree of freedom while excluding cases and households in the *undefined* SES level. As a sensitivity analysis, we included cases and households in the *undefined* level, grouping them with SES levels *D* and *E*. All statistical analyses were performed in R version 4.2.1.^40^ The Cochran-Armitage trend tests were performed using the *CochranArmitageTest* function in the *DescTools* package, and chi-squared tests were performed using the *chisq.test* function in the base *stats* package.^40,44^

### Assessing the impact of active surveillance on observed disparities

To understand how the supplemental active surveillance activities that began in 2021 impacted rabies disparities, we assessed the SES distributions of samples obtained via active vs. passive surveillance in 2021 and 2022 and tested for differences using chi-squared tests. We compared plots of the temporal trends in the number of cases and submitted samples by SES from the main results to similar plots that excluded samples obtained via active surveillance. We repeated our analysis of the trend between sample positivity and neighborhood SES but excluding samples obtained via active surveillance. We repeated the one-sided Cochran-Armitage test excluding active samples for the 2021-2022 period (the only period with active surveillance) as well as the entire study period and compared these results to the main results that were calculated using all (both active and passive) samples.

### Spatial analysis of sample positivity

We used a generalized additive model (GAM) to determine how sample positivity varied spatially and assess for the presence of global spatial heterogeneity and local hotspots of sample positivity. Spatial analysis using GAM has been described previously,^45,46^ but in brief, the GAM was used to generate an odds ratio map of sample positivity across Arequipa. A global test was performed to test for overall spatial heterogeneity with an alpha of 0.05, and given a significant global test, a local test for significance was performed to identify areas (hotspots) with higher sample positivity using a one-sided alpha of 0.01. We considered both a crude GAM without covariate adjustment and a GAM that included adjustment for locality-level SES and distance between the locality centroid of the sample of origin and the nearest health facility. Spatial analyses were performed using R package *MapGAM*,^40,47^ and risk maps generated from model outputs were created using R package *ggmap*.^43^

## RESULTS

### Rabies cases and samples

There were 347 confirmed cases of dog rabies in 2015-2022 (Figure 1a). Of these 347 cases, only two cases (0.6%) were located >50 m from the closest city block and were thus excluded from further analyses. Of the remaining 345 cases, 203 (58.8%) were located inside a city block, while 142 (41.2%) fell within 0.1-46 m of the closest city block. Six (1.7%) were assigned to SES level A, 21 (6.1%) to level B, 46 (13.3%) to level C, 120 (34.6%) to level D, 129 (37.2%) to level E, and 23 (6.6%) to level *undefined*. Compared to the SES distribution of households in the city, cases were disproportionately concentrated in disadvantaged city blocks, i.e., SES levels *D* and *E* (*p* < 0.0001; Figure S1 and Table S1). These results were unchanged when *undefined* cases and households were included in the analysis grouped with SES levels *D* and *E* (*p* < 0.0001).

**Figure 1.**
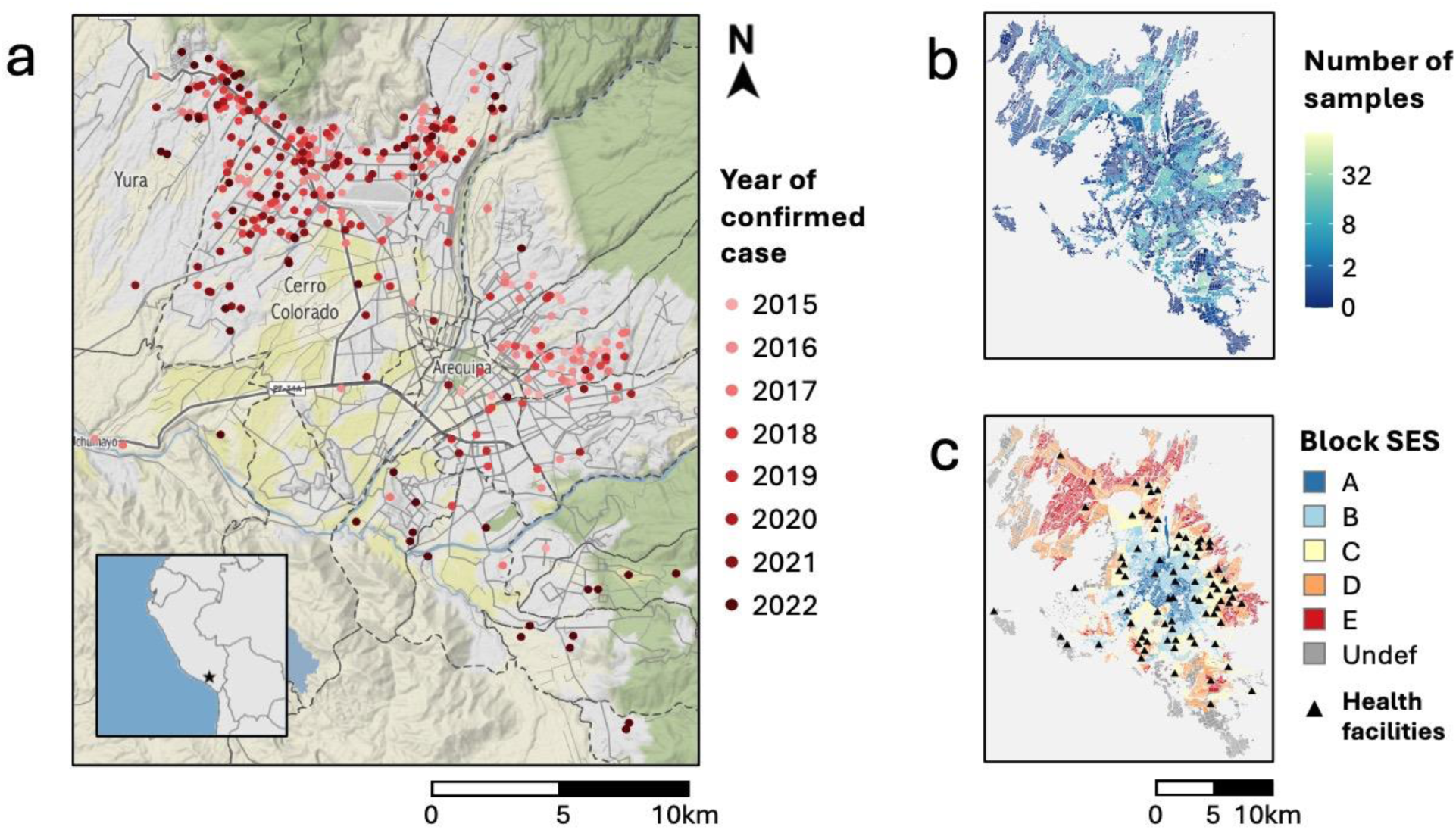
Maps depicting the confirmed cases of canine rabies (a), surveillance effort by locality (b), and block-level socioeconomic status (c). Panel a shows the locations of confirmed cases of canine rabies as dots colored according to year. Case locations were slightly jittered to protect the privacy of positive households. District boundaries are indicated by dashed lines, and the districts of Yura and Cerro Colorado are labeled. The inset map in panel a shows the location of Arequipa in Peru. Panel b maps the number of samples submitted for laboratory testing for canine rabies in 2015-2022 for each locality of Arequipa, and the color scale was log(*x*+1) transformed. Panel c depicts the geographic distribution of block-level socioeconomic status (SES) in Arequipa and the locations of health facilities (black triangles). Block-level SES is an ordinal variable, with group A comprising the most affluent blocks and group E comprising the most disadvantaged blocks. Blocks whose SES level was undefined (“Undef”) are colored gray. Note that the spatial extent of all three map panels are the same.

In the same eight-year span, there were 2,568 samples submitted for laboratory testing of rabies virus. Of these, 2,119 (82.5%) samples could be matched to a locality of origin; the 449 (17.5%) samples missing locality information were excluded from further analyses. Of the 2,119 samples that could be matched to a locality of origin, 107 (5%) originated from localities in the highest SES level (A), 352 (16.6%) were from localities in level B, 614 (29%) were from localities in level C, 583 (27.5%) were from localities in level D, 431 (20.3%) were from localities in level E, and 32 (1.5%) were from localities in the *undefined* SES level.

### Temporal trends and sample positivity

During the initial years of the epidemic, rabies cases were typically detected just east of the city center; however, cases have increasingly extended into the northwestern and southern periphery in recent years (Figure 1a). Surveillance effort has been spatially heterogeneous, with greater sample submission volumes in localities closer to the city center, particularly in the east and south (Figure 1b). Health facilities in Arequipa are concentrated in the more affluent neighborhoods located in or near the city center (Figure 1c). Although many recent cases have been detected in the disadvantaged peri-urban region of the northwest, sample volume has been relatively low in that area, which also has scarce health facilities (Figure 1a-c).

Since the second year of the epidemic (2016), there has been a clear and sustained disparity in the number of confirmed rabies cases in which annual case counts have been highest in the most disadvantaged areas (*D* and *E*; Figure 2a). Surveillance effort - which was highest at the beginning of the epidemic and dipped in 2020 during the first year of the COVID-19 pandemic - shows a somewhat different temporal trend; annual sample counts were initially highest in localities with intermediate SES, but submissions from disadvantaged localities have made up a greater share of annual surveillance effort over time (Figure 2b).

**Figure 2.**
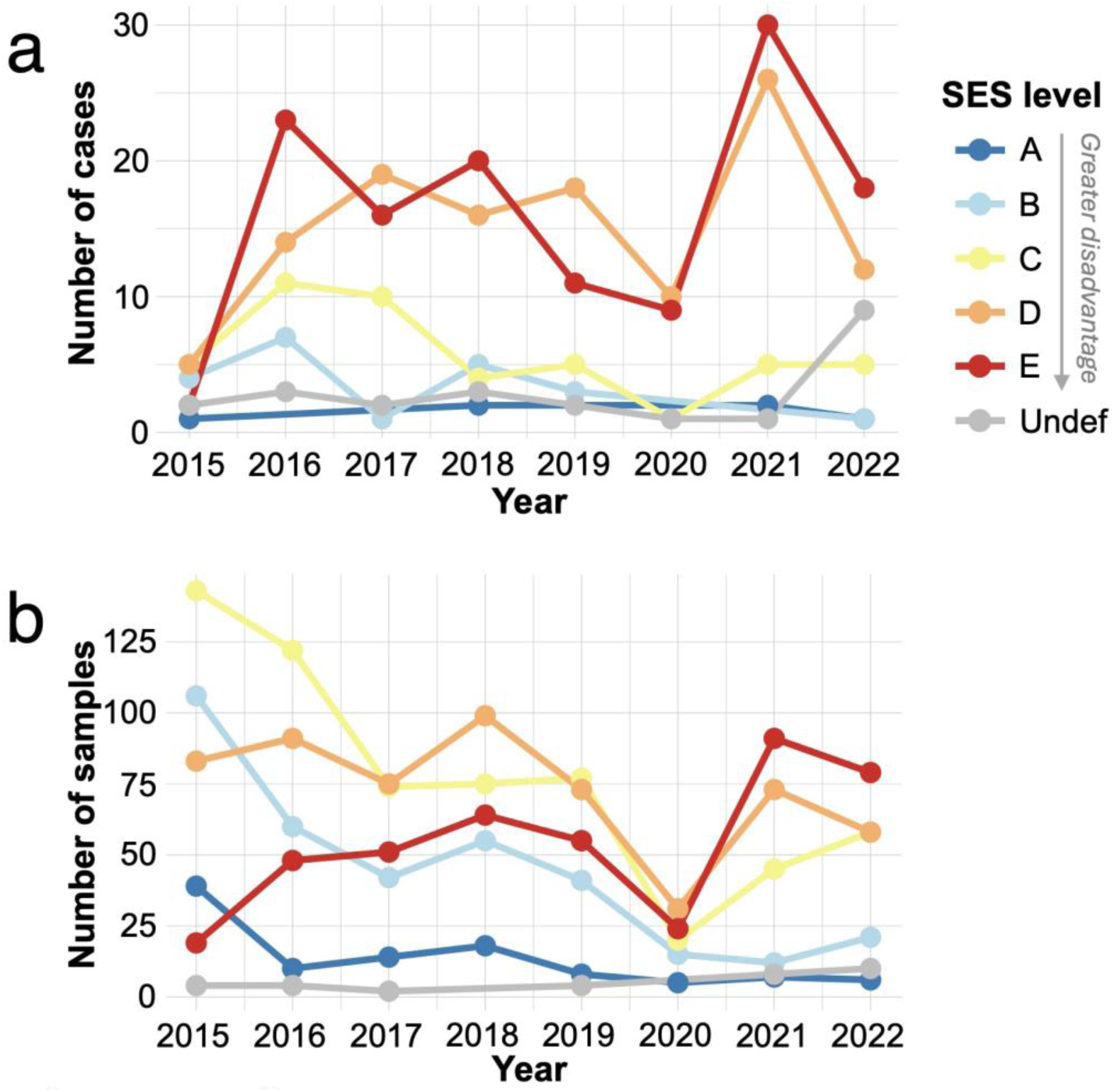
Temporal trends in the number of confirmed cases (a) and submitted samples (b) by socioeconomic status. Lines are colored according to the socioeconomic (SES) level assigned to the case or sample of origin, where cases were assigned to SES at the block level, and samples were assigned to SES at the locality level. SES is an ordinal variable ranging *A*-*E*, with *E* denoting the most disadvantaged level.

Sample positivity increased with neighborhood deprivation, and this trend was highly significant across all periods considered (*p* < 0.0001 across all years and for each two-year interval except for 2021-2022, which had *p* = 0.012; Figure 3). The inclusion of *undefined* blocks (reassigned to level *E*) did not change the significance of the association for all periods considered (*p* = 0.013 for 2021-22 and *p <* 0.0001 for all other periods). The period with the weakest trend between sample positivity and neighborhood SES (2021-2022) coincided with supplemental active surveillance activities. In the next section, we discuss how these activities may have impacted observed trends.

**Figure 3.**
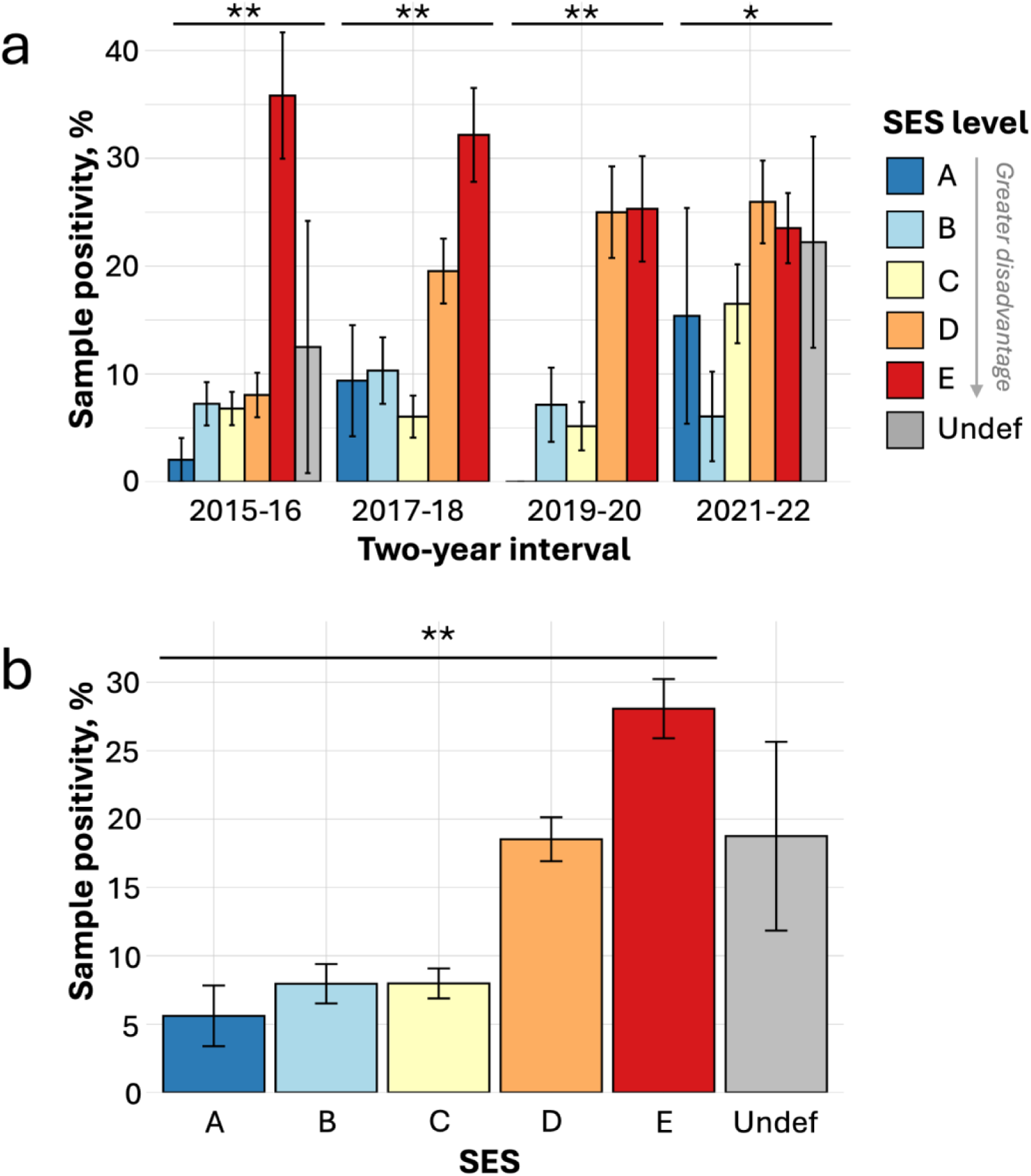
Sample positivity by socioeconomic status (SES) level for canine rabies samples plotted for two-year intervals (a) and across all years (b). Bars are plotted with their margins of error and shaded according to the SES level associated with the sample of origin. SES is an ordinal variable ranging *A*-*E*, with *E* denoting the most disadvantaged level. There were <5 samples from *undefined* blocks in 2017-18 and 2019-20, and these bars were not plotted, as they had very large margins of error. The results of Cochran-Armitage trend tests for a positive association between sample positivity and neighborhood disadvantage (SES A → E) are indicated by asterisks: ** denotes *p* <0.001 and * denotes *p* < 0.05.

### Impacts of active surveillance on SES disparities

Of the 2,119 samples submitted for rabies testing, 153 were obtained via supplemental active surveillance in 2021-2022 (Supplementary Table S2). Active surveillance targeted dry water channels, a strategy informed by eco-epidemiological data suggesting that these landscape features may act as a conduit for rabies transmission among the local dog population.^29,35^ While active surveillance was not intentionally aimed at socioeconomically disadvantaged areas, dry water channels are more concentrated in disadvantaged areas, and samples collected from active surveillance activities were more likely to be from disadvantaged localities (i.e., those assigned to SES levels *D*, *E*, or *undefined*) compared to samples submitted via passive surveillance (*p* = 0.017 and 0.009 for 2021 and 2022, respectively). While samples from disadvantaged localities accounted for 67% and 58% of passive samples in 2021 and 2022, respectively, they accounted for 81% and 78% of active samples during the same years (Figure S2). Only a small number of positive samples were collected via active surveillance (5 positives out of 153 active samples compared to 105 of 315 passive samples), and the inclusion of actively collected samples had minimal impact on temporal trends in case numbers across SES levels (compare Figure 2a to Figure S3a). However, the inclusion of samples collected via active surveillance markedly increased the surveillance effort in the most disadvantaged neighborhoods (compare Figure 2b to Figure S3b).

Compared to all samples submitted in 2021-2022 (both active and passive), passive samples from 2021 have more striking disparities in sample positivity, which was more strongly associated with neighborhood disadvantage (*p* < 0.0001 compared to *p* = 0.012 for all 2021-2022 samples; Figure S4a). These trends were also apparent across all years: sample positivity in high and intermediate SES groups was unchanged when comparing all samples to passive samples only, but sample positivity was higher in low SES groups when active samples were excluded (compare Figure 3b to Figure S4b). These results indicate that active surveillance contributed a greater share of samples from disadvantaged localities compared to passive surveillance and reduced disparities in both surveillance effort and sample positivity.

### Spatial analysis results

Spatial modeling using GAM revealed that sample positivity varied across Arequipa (global test *p* < 0.001; Figure 4). Local one-sided significance tests, used to identify areas where the odds of samples testing positive were significantly higher than the overall mean, detected a hotspot of sample positivity in the northwest comprising Cerro Colorado and Yura districts (*p* < 0.01; Figure 4). This area is marked by socioeconomic disadvantage and low access to health facilities (Figure 1c). Adjusting for SES and distance from the locality centroid to the closest health facility reduced the intensity and size of the hotspot, though it persisted in the adjusted model (Figure 4b).

**Figure 4.**
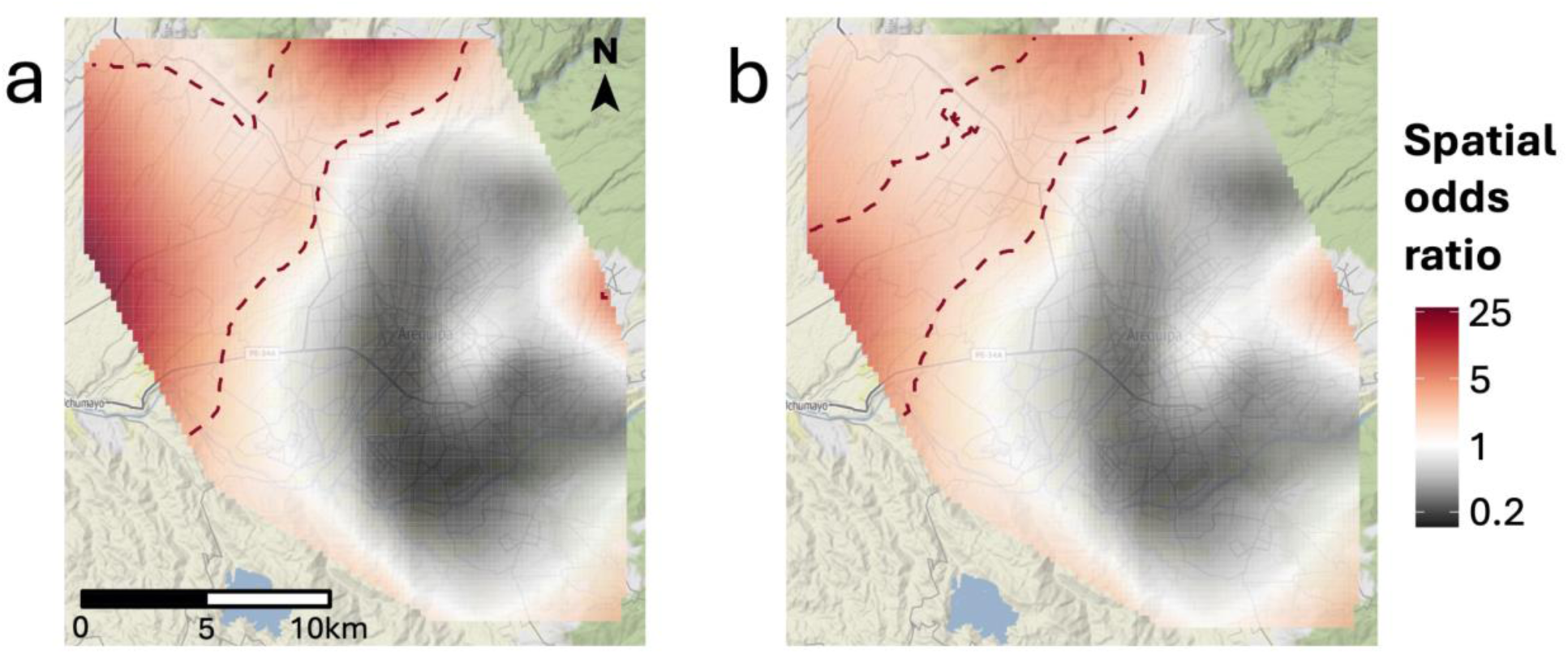
The spatial risk of sample positivity before (a) and after (b) covariate adjustment for neighborhood socioeconomic status and distance to the nearest health facility. Raster cells are colored according to the spatial odds ratio of sample positivity, with red denoting areas of increased risk. Statistically significant hotspots (*p* < 0.01) of sample positivity are demarcated by red, dotted lines.

## DISCUSSION

While dog-mediated rabies is commonly linked to poverty, few studies have investigated how dog rabies incidence varies with neighborhood SES. Our analysis revealed clear socioeconomic disparities in dog rabies burden in Arequipa, Peru: confirmed cases and sample positivity rates were highest in disadvantaged neighborhoods. Additionally, our spatial analysis identified a sample positivity hotspot in the peri-urban region located northwest of the city center, which could be explained, in part, by the disadvantage and low access to health facilities in that area. As sample positivity provides insight into both case incidence and surveillance effort, the disparities in this metric also suggest that surveillance effort has been insufficient in disadvantaged areas of the city. Encouragingly, our results also indicate that spatially-targeted active surveillance can be combined with population-based passive surveillance to mitigate disparities in surveillance effort and sample positivity.

Our study adds to the evidence that the risk of dog-mediated rabies is disproportionately higher in the most disadvantaged areas of Arequipa. Previous studies of dog-mediated rabies risk in the city have found much higher rates of dog bites in the city’s peri-urban areas.^17^ Once bitten, the percentage of people who received medical care including PEP in the city’s wealthier urban areas was almost twice as high compared to those living in the city’s peri-urban areas.^17^ Indeed, the first, and so far only, documented human fatality due to rabies in the region occurred in Chiguata, a peri-urban area east of the city center.^48^ The low access and utilization of health services in disadvantaged peri-urban areas likely contributes to underdetection of human cases. Studies assessing local knowledge, attitude, and practices towards rabies conducted in other LMICs have found socioeconomic disadvantage to be associated with less knowledge about rabies preventative practices and treatment,^18–25^ a mechanism by which socioeconomic disadvantage may also act as a social determinant of human rabies in Arequipa.

Canine rabies incidence is higher in Arequipa’s disadvantaged neighborhoods due to several factors. Although vaccination coverage is similar across SES levels, peri-urban areas have more free-roaming dogs, which experience higher contact rates and population turnover, reducing effective vaccination coverage.^49^ The greater concentration of free-roaming dogs in peri-urban areas is partly due to inadequate waste management policies and budgets compared to urban areas, resulting in greater accumulations of solid waste, an important resource for free-roaming dogs.^45^ Arequipa’s unique geographical features, dry water channels, have been shown to allow free-roaming dogs to travel long distances, increasing transmission potential.^35^ These water channels have more access points in the peri-urban areas compared to urban central areas. Additionally, peri-urban dogs are often bred as guard dogs, leading to more aggressive behavior over time.^30^ Taken together with past research, our findings suggest that the unique demographics of peri-urban dogs contribute to the disproportionate number of rabid dogs in disadvantaged areas.

The disparity in sample positivity and the lesser relative surveillance effort in disadvantaged areas can be explained by a variety of factors. First, the passive surveillance system requires that residents report dead or suspected dogs to nearby health facilities. However, disadvantaged, peri-urban parts of the city have relatively low access to health facilities, making reporting more challenging in these areas. In fact, the hot spot of sample positivity we detected was located in the city’s northwestern peri-urban periphery. Our analysis also revealed that this same area has the lowest density of health facilities, and the adjusted spatial GAM model indicated that distance to health facilities could explain some of the increased positivity in that area. In previous work, distance to health facilities has been associated with other rabies outcomes, including decreased PEP uptake and greater number of deaths due to rabies.^17,24,50^ In addition to decreased health access, those living in peri-urban areas work informal jobs that preclude them from going to health posts to report suspicious dogs. Our findings are in line with previous work suggesting evidence of under-reporting of dog bites in peri-urban areas (a related kind of surveillance).^17^ Finally, reporting of suspicious dogs requires that residents be informed about rabies, and knowledge of rabies may be poorer in disadvantaged, peri-urban areas.

Our study is subject to some limitations. First, our analyses relied on using data from a predominantly passive surveillance system, which is subject to reporting bias, including differential reporting by SES (as health service access and knowledge of rabies is likely lower in disadvantaged areas). However, as explained below, including negative samples allowed us to account for differential surveillance effort. Another limitation is the 17.5% of samples that were excluded from our analysis due to missing locality information. Additionally, the locations of negative samples were only available at the locality level, which may have resulted in some misclassification of sample SES. However, due to the high spatial correlation of block-level SES, there was good agreement between block- and locality-level SES (Cohen’s k = 0.834), suggesting that misclassification was low. Finally, using locality centroids to represent the locations of negative samples in the spatial GAM analysis resulted in some inaccuracy in the control locations. However, localities are relatively small geographic units; there are 1,317 localities in Arequipa and each contains a median of only 13 city blocks. Thus, the degree of spatial misclassification introduced by using centroids at this level of spatial aggregation likely had minimal impact on our results.

A key strength of our study is the inclusion of both positive and negative samples, allowing us to account for variation in surveillance effort across space and SES. Previous studies relying only on confirmed cases^27,28^ may misidentify high-risk areas, as high case counts often reflect greater surveillance effort. For example, a study in El Salvador reported higher dog rabies incidence in low-poverty areas, likely influenced by underreporting in high-poverty rural regions.^28^ By combining case incidence with sample positivity, we provide a more nuanced view of rabies disparities. Our finding that disadvantaged areas have both the highest case counts and sample positivity suggests true disparities may be even greater than case data alone indicate. Using high-spatial-resolution data enabled us to examine SES impacts within communities in a single city, unlike prior studies conducted on broader spatial scales.^10,11,27,28^

In conclusion, we found that global trends in rabies disparities were also evident on a local scale, which highlight a critical health equity issue in Latin America. Disadvantaged areas are disproportionately affected by dog rabies and should be prioritized to protect the most vulnerable. By identifying where resources are most needed, our analysis supports a shift from broad city-wide approaches, necessary in some contexts (i.e., city-wide mass dog vaccination), to more targeted surveillance and control strategies that use limited resources efficiently. In addition to reducing inequities, targeting resources to the highest-risk areas would support rabies elimination, as spatial heterogeneity in transmission risk can sustain an epidemic even when herd immunity thresholds have been reached.^51,52^ Importantly, our results advance the goals of One Health by integrating human, animal, and environmental health. By focusing on socioeconomic and geographic inequities, our study emphasizes the need for coordinated efforts across multiple sectors, including healthcare, veterinary services, urban planning, and local stakeholder engagement. Reducing inequities in dog rabies control will be essential for achieving the WHO’s goal of eliminating dog-mediated human rabies by 2030.^53^

## Data Availability

All code and non-sensitive data are available on Github: https://github.com/RabiesLabPeru/ses_rabies.

## AUTHOR CONTRIBUTIONS

SX contributed to the conceptualization, data curation, formal analysis, methodology, visualization, and writing of the original manuscript. JS contributed to writing the original manuscript and reviewing and editing the final manuscript. ED, EZ, and YM contributed to the investigation and project administration. SR contributed to writing of the paper and reviewing the final manuscript RCN contributed to the conceptualization, data curation, formal analysis, funding acquisition, investigation, methodology, project administration, supervision, and writing of the paper.

## REFERENCES

1. Hampson K, Coudeville L, Lembo T, Sambo M, Kieffer A, Attlan M, et al. Estimating the Global Burden of Endemic Canine Rabies. PLOS Neglected Tropical Diseases. 2015 Apr 16;9(4):e0003709.

2. Gan H, Hou X, Wang Y, Xu G, Huang Z, Zhang T, et al. Global burden of rabies in 204 countries and territories, from 1990 to 2019: results from the Global Burden of Disease Study 2019. International Journal of Infectious Diseases. 2023 Jan 1;126:136–44.

3. Scott TP, Nel LH. Lyssaviruses and the Fatal Encephalitic Disease Rabies. Front Immunol [Internet]. 2021 Dec 2 [cited 2024 Sep 4];12. Available from: https://www.frontiersin.org/journals/immunology/articles/10.3389/fimmu.2021.786953/full

4. Bucher A, Dimov A, Fink G, Chitnis N, Bonfoh B, Zinsstag J. Benefit-cost analysis of coordinated strategies for control of rabies in Africa. Nat Commun. 2023 Sep 7;14(1):5370.

5. World Health Organization. WHO Fact Sheet on Rabies [Internet]. [cited 2024 Nov 13]. Available from: https://www.who.int/news-room/fact-sheets/detail/rabies

6. Seetahal JFR, Vokaty A, Vigilato MAN, Carrington CVF, Pradel J, Louison B, et al. Rabies in the Caribbean: A Situational Analysis and Historic Review. Tropical Medicine and Infectious Disease. 2018 Sep;3(3):89.

7. Meske M, Fanelli A, Rocha F, Awada L, Soto PC, Mapitse N, et al. Evolution of Rabies in South America and Inter-Species Dynamics (2009–2018). Tropical Medicine and Infectious Disease. 2021 Jun;6(2):98.

8. Vigilato MAN, Clavijo A, Knobl T, Silva HMT, Cosivi O, Schneider MC, et al. Progress towards eliminating canine rabies: policies and perspectives from Latin America and the Caribbean. Philos Trans R Soc Lond B Biol Sci. 2013 Aug 5;368(1623):20120143.

9. World Health Organization. WHO expert consultation on rabies: third report [Internet]. Geneva: World Health Organization; 2018 [cited 2022 Sep 21]. 183 p. (WHO technical report series;1012). Available from: https://apps.who.int/iris/handle/10665/272364

10. Taylor E, George K, Johnson E, Whitelegg H, Prada JM, Horton DL. Quantifying the interconnectedness between poverty, health access, and rabies mortality. PLoS neglected tropical diseases. 2023;17(4):e0011204.

11. Schneider MC, Aguilera XP, Barbosa Da Silva Junior J, Ault SK, Najera P, Martinez J, et al. Elimination of Neglected Diseases in Latin America and the Caribbean: A Mapping of Selected Diseases. Brooker S, editor. PLoS Negl Trop Dis. 2011 Feb 15;5(2):e964.

12. Alemayehu T, Oguttu B, Rupprecht CE, Niyas VKM. Rabies vaccinations save lives but where are the vaccines? Global vaccine inequity and escalating rabies-related mortality in low-and middle-income countries. International Journal of Infectious Diseases. 2024;140:49–51.

13. Salahuddin N, Blumberg L, Abela B, Durrheim DN. GAVI investment should accelerate Rabies “Zero by 30” aspiration. IJID One Health. 2024 Dec 1;5:100045.

14. Wambura G, Mwatondo A, Muturi M, Nasimiyu C, Wentworth D, Hampson K, et al. Rabies vaccine and immunoglobulin supply and logistics: challenges and opportunities for rabies elimination in Kenya. Vaccine. 2019;37:A28–34.

15. Baron JN, Chevalier V, Ly S, Duong V, Dussart P, Fontenille D, et al. Accessibility to rabies centers and human rabies post-exposure prophylaxis rates in Cambodia: A Bayesian spatio-temporal analysis to identify optimal locations for future centers. PLoS Neglected Tropical Diseases. 2022;16(6):e0010494.

16. Castillo-Neyra R, Buttenheim AM, Brown J, Ferrara JF, Arevalo-Nieto C, Borrini-Mayorí K, et al. Behavioral and structural barriers to accessing human post-exposure prophylaxis and other preventive practices in Arequipa, Peru, during a canine rabies epidemic. PLOS Neglected Tropical Diseases. 2020 Jul 21;14(7):e0008478.

17. De la Puente-León M, Levy MZ, Toledo AM, Recuenco S, Shinnick J, Castillo-Neyra R. Spatial Inequality Hides the Burden of Dog Bites and the Risk of Dog-Mediated Human Rabies. Am J Trop Med Hyg. 2020 Sep;103(3):1247–57.

18. Tiwari HK, Robertson ID, O’Dea M, Vanak AT. Knowledge, attitudes and practices (KAP) towards rabies and free roaming dogs (FRD) in Panchkula district of north India: A cross-sectional study of urban residents. PLoS Negl Trop Dis. 2019 Apr 29;13(4):e0007384.

19. Sambo M, Lembo T, Cleaveland S, Ferguson HM, Sikana L, Simon C, et al. Knowledge, Attitudes and Practices (KAP) about Rabies Prevention and Control: A Community Survey in Tanzania. PLoS Negl Trop Dis. 2014 Dec 4;8(12):e3310.

20. Pal P, Yawongsa A, Bhusal TN, Bashyal R, Rukkwamsuk T. Knowledge, attitude, and practice about rabies prevention and control: A community survey in Nepal. Vet World. 2021 Apr;14(4):933–42.

21. Hagos WG, Muchie KF, Gebru GG, Mezgebe GG, Reda KA, Dachew BA. Assessment of knowledge, attitude and practice towards rabies and associated factors among household heads in Mekelle city, Ethiopia. BMC Public Health. 2020 Jan 14;20(1):57.

22. Costa GB, Gilbert A, Monroe B, Blanton J, Ngam SN, Recuenco S, et al. The influence of poverty and rabies knowledge on healthcare seeking behaviors and dog ownership, Cameroon. PLOS ONE. 2018 Jun 21;13(6):e0197330.

23. Fang LX, Ping F, Hui BG, Yan YX. Socioeconomic status is a critical risk factor for human rabies post-exposure prophylaxis. Vaccine. 2010;28(42):6847–51.

24. Hampson K, Dobson A, Kaare M, Dushoff J, Magoto M, Sindoya E, et al. Rabies exposures, post-exposure prophylaxis and deaths in a region of endemic canine rabies. PLoS neglected tropical diseases. 2008;2(11):e339.

25. Kisaka S, Makumbi F, Majalija S, Bahizi G, Thumbi S. Delays in initiating rabies post-exposure prophylaxis among dog bite victims in Wakiso and Kampala districts, Uganda. AAS Open Res. 2022 Dec 8;4:49.

26. Wallace RM, Mehal J, Nakazawa Y, Recuenco S, Bakamutumaho B, Osinubi M, et al. The impact of poverty on dog ownership and access to canine rabies vaccination: results from a knowledge, attitudes and practices survey, Uganda 2013. Infectious Diseases of Poverty. 2017 Jun 1;6(1):97.

27. Kanankege KST, Errecaborde KM, Wiratsudakul A, Wongnak P, Yoopatthanawong C, Thanapongtharm W, et al. Identifying high-risk areas for dog-mediated rabies using Bayesian spatial regression. One Health. 2022 Dec 1;15:100411.

28. Arias-Orozco P, Bástida-González F, Cruz L, Villatoro J, Espinoza E, Zárate-Segura PB, et al. Spatiotemporal analysis of canine rabies in El Salvador: Violence and poverty as social factors of canine rabies. Rupprecht CE, editor. PLoS ONE. 2018 Aug 17;13(8):e0201305.

29. Castillo-Neyra R, Zegarra E, Monroy Y, Bernedo R, Cornejo-Rosello I, Paz-Soldan V, et al. Spatial Association of Canine Rabies Outbreak and Ecological Urban Corridors, Arequipa, Peru. TropicalMed. 2017 Aug 13;2(3):38.

30. Castillo-Neyra R, Brown J, Borrini K, Arevalo C, Levy MZ, Buttenheim A, et al. Barriers to dog rabies vaccination during an urban rabies outbreak: Qualitative findings from Arequipa, Peru. Recuenco S, editor. PLoS Negl Trop Dis. 2017 Mar 17;11(3):e0005460.

31. Castillo-Neyra R, Toledo AM, Arevalo-Nieto C, MacDonald H, De la Puente-León M, Naquira-Velarde C, et al. Socio-spatial heterogeneity in participation in mass dog rabies vaccination campaigns, Arequipa, Peru. Blanton J, editor. PLoS Negl Trop Dis. 2019 Aug 1;13(8):e0007600.

32. Castillo-Neyra R, Xie S, Bellotti BR, Diaz EW, Saxena A, Toledo AM, et al. Optimizing the location of vaccination sites to stop a zoonotic epidemic. Sci Rep. 2024 Jul 10;14(1):15910.

33. Xie S, Rieders M, Changolkar S, Bhattacharya BB, Diaz EW, Levy MZ, et al. Enhancing mass vaccination programs with queueing theory and spatial optimization. Front Public Health. 2024 Dec 24;12.

34. Instituto Nacional de Salud. Manual de procedimientos para el diagnóstico de la rabia [Internet]. Lima, Perú: Ministerio de Salud; 2002 [cited 2024 Oct 3]. Available from: https://bvs.minsa.gob.pe/local/MINSA/1047_INS-NT31.pdf

35. Raynor B, De la Puente-León M, Johnson A, Díaz EW, Levy MZ, Recuenco SE, et al. Movement patterns of free-roaming dogs on heterogeneous urban landscapes: Implications for rabies control. Preventive Veterinary Medicine. 2020 May;178:104978.

36. Carhuavilca Bonnett D, Sánchez Aguilar A. Planos Estratificados de Lima Metropolitana a Nivel de Manzanas 2020. Instituto Nacional de Estadística e Informática [Internet]. 2020 [cited 2024 Dec 1]; Available from: https://www.inei.gob.pe/media/MenuRecursivo/publicaciones_digitales/Est/Lib1744/libro.pdf

37. Elbers C, Lanjouw JO, Lanjouw P. Micro-Level Estimation of Poverty and Inequality. Econometrica. 2003 Jan;71(1):355–64.

38. Dalenius T, and Hodges Jr JL. Minimum Variance Stratification. Journal of the American Statistical Association. 1959 Mar 1;54(285):88–101.

39. Levy MZ, Barbu CM, Castillo-Neyra R, Quispe-Machaca VR, Ancca-Juarez J, Escalante-Mejia P, et al. Urbanization, land tenure security and vector-borne Chagas disease. Proceedings of the Royal Society B: Biological Sciences. 2014 Aug 22;281(1789):20141003.

40. R Core Team. R: A language and environment for statistical computing [Internet]. Vienna, Austria: R Foundation for Statistical Computing; 2024. Available from: https://www.R-project.org

41. Pebesma E, Bivand R. Spatial Data Science: With applications in R [Internet]. Chapman and Hall/CRC; Available from: https://r-spatial.org/book/

42. Pebesma E. Simple Features for R: Standardized Support for Spatial Vector Data. The R Journal. 2018;10(1):439–46.

43. Kahle D, Wickham H. ggmap: Spatial Visualization with ggplot2. The R Journal. 2013;5(1):144–61.

44. Signorell A, Aho K, Alfons A, Anderegg N, Aragon T, Arachchige C, et al. DescTools: Tools for Descriptive Statistics [Internet]. 2024 [cited 2024 Jun 10]. Available from: https://cran.r-project.org/web/packages/DescTools/index.html

45. Xie S, Hubbard RA, Himes BE. Analysis of Spatial Trends in Smoking Status Among Patients with Obstructive Airway Diseases Highlight Potential for Targeted Interventions. AMIA Annual Symposium Proceedings. 2019;

46. Webster T, Vieira V, Weinberg J, Aschengrau A. Method for mapping population-based case-control studies: an application using generalized additive models. Int J Health Geogr. 2006;5:26.

47. Bai L, Bartell S, Vieira V. MapGAM: Mapping Smoothed Effect Estimates from Individual-Level Data [Internet]. 2023. Available from: https://CRAN.R-project.org/package=MapGAM

48. Gómez Vega R. Una mujer que fue mordida por un perro callejero muere de rabia en Arequipa [Internet]. El País. 2023 [cited 2024 Jul 15]. Available from: https://elpais.com/sociedad/2023-10-18/una-mujer-que-fue-mordida-por-un-perro-callejero-muere-de-rabia-en-arequipa.html

49. Chuquista-Alcarraz O, Falcón N, Vigilato MAN, Rocha F, Toledo-Barone G, Amorim-Conselheiro J, et al. Dog Population Rabies Immunity before a Mass Vaccination Campaign in Lima, Peru: Vulnerabilities for Virus Reestablishment. The American Journal of Tropical Medicine and Hygiene. 2023 Jul 10;109(2):420.

50. Joseph J, Sangeetha N, Khan AM, Rajoura OP. Determinants of delay in initiating post-exposure prophylaxis for rabies prevention among animal bite cases: hospital based study. Vaccine. 2013;32(1):74–7.

51. Hampson K, Dushoff J, Cleaveland S, Haydon DT, Kaare M, Packer C, et al. Transmission Dynamics and Prospects for the Elimination of Canine Rabies. Rupprecht CE, editor. PLoS Biol. 2009 Mar 10;7(3):e1000053.

52. Townsend SE, Lembo T, Cleaveland S, Meslin FX, Miranda ME, Putra AAG, et al. Surveillance guidelines for disease elimination: A case study of canine rabies. Comparative Immunology, Microbiology and Infectious Diseases. 2013 May 1;36(3):249– 61.

53. World Health Organization, Food and Agricultural Organization of the United Nations, World Organisation for Animal Health, Global Alliance for Rabies Control. Zero by 30: The Global Strategic Pland to End Human Deaths from Dog-Mediated Rabies by 2030 [Internet]. Geneva; 2018 [cited 2024 Nov 18]. Available from: https://iris.who.int/bitstream/handle/10665/272756/9789241513838-eng.pdf

