## Supplementary materials for "Neighborhood Socioeconomic Status and Dog-Mediated Rabies: Disparities in Incidence and Surveillance Effort in a Latin American City"

### Contents

**Supplementary Table S1.** The socioeconomic status distributions (SES) of rabies cases, submitted samples, and geographic units used in the analysis ………………………………………………………. 2

**Supplementary Table S1.** Number of canine samples collected via active and passive surveillance for rabies testing in Arequipa, Peru by year…………………………………………………………………... 3

**Figure S1.** The socioeconomic distribution of dog rabies cases compared to households in Arequipa, Peru. 4

**Supplementary Table S1. The socioeconomic status distributions (SES) of rabies cases, submitted samples, and geographic units used in the analysis.** Numbers shown are counts with percentages shown in parentheses.

| SES level | Cases, *n* (%) | Samples, *n* (%) | Houses, *n* (%) | Blocks, *n* (%) | Localities, *n* (%) |
|---|---|---|---|---|---|
| **A** | 6 (1.7) | 107 (5.0) | 15,369 (9.1) | 1,418 (5.2) | 130 (9.9) |
| **B** | 21 (6.1) | 352 (16.6) | 32,289 (19.1) | 2,961 (10.9) | 181 (13.7) |
| **C** | 46 (13.3) | 614 (29.0) | 39,843 (23.5) | 4,596 (16.9) | 254 (19.3) |
| **D** | 120 (34.8) | 583 (27.5) | 39,729 (23.4) | 7,238 (26.6) | 354 (26.9) |
| **E** | 129 (37.4) | 431 (20.3) | 36,242 (21.4) | 5,834 (21.4) | 243 (18.5) |
| **Undef** | 23 (6.7) | 32 (1.5) | 5,955 (3.5) | 5,171 (19.0) | 155 (11.8) |
| **Total** | 345 (100) | 2,119 (100) | 169,427 (100) | 27,218 (100) | 1,317 (100) |

**Supplementary Table S2. Number of canine samples collected via active and passive surveillance for rabies testing in Arequipa, Peru by year.** Counts of positive samples for each year and surveillance mechanism are given in parentheses.

| | Yearly total sample count (positive sample count) | | | | | | | | |
|---|---|---|---|---|---|---|---|---|---|
| | **2015** | **2016** | **2017** | **2018** | **2019** | **2020** | **2021** | **2022** | **Total** |
| **Passive** | 394 (19) | 335 (58) | 258 (48) | 311 (50) | 258 (39) | 95 (21) | 146 (59) | 169 (46) | 1,966 (340) |
| **Active** | 0 | 0 | 0 | 0 | 0 | 0 | 90 (5) | 63 (0) | 153 (5) |
| **Total** | 394 (19) | 335 (58) | 258 (48) | 311 (50) | 258 (39) | 95 (21) | 236 (64) | 232 (46) | 2,119 (345) |

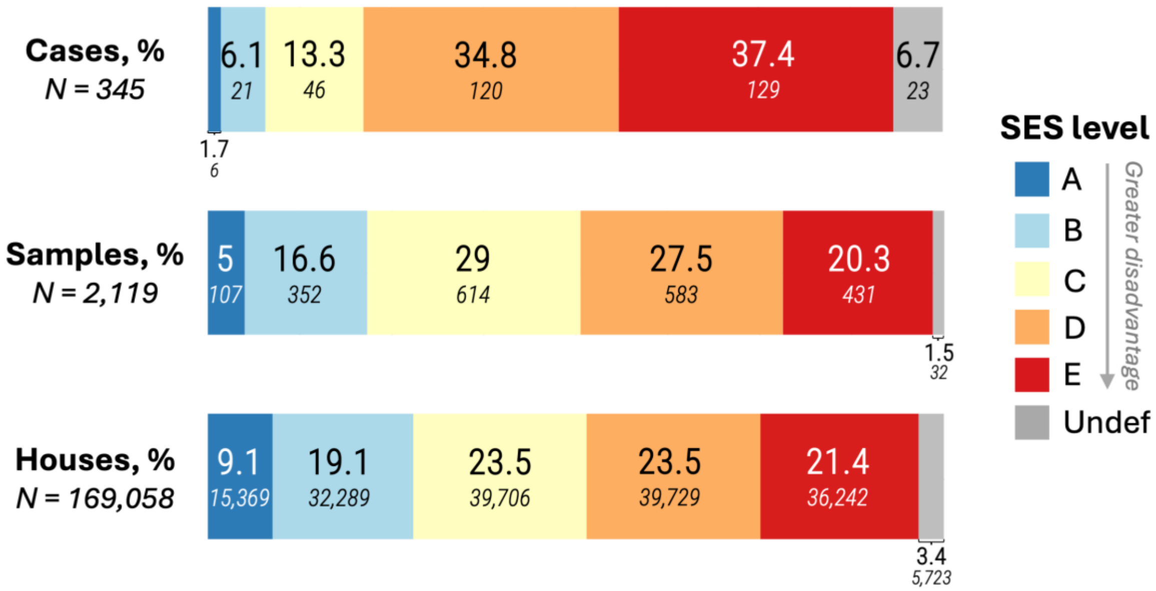

**Figure S1. The socioeconomic distribution of dog rabies cases and submitted samples compared to households in Arequipa, Peru.** Stacked row charts show the proportion of cases, samples, and households falling within each SES level. Large numbers indicate the percentage (%) distribution and the small number in italics give the count of cases, samples, or houses falling in each category.

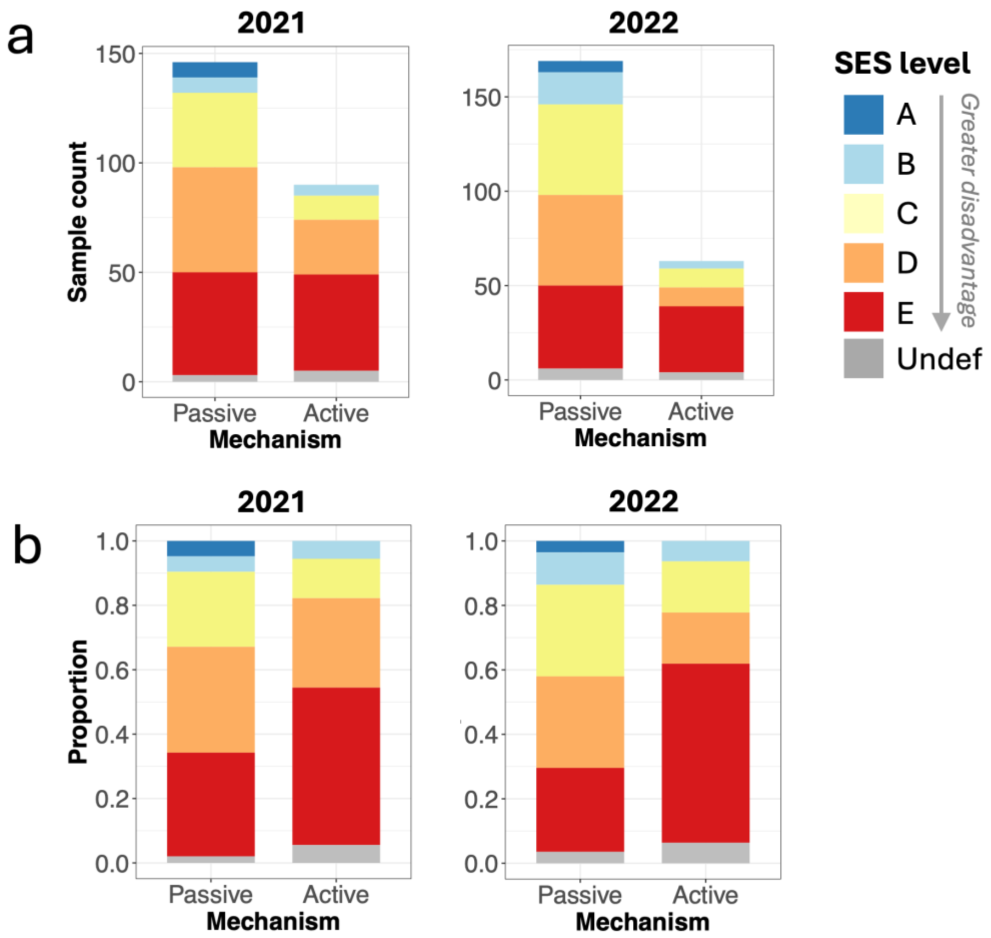

**Figure S2. Comparison of the socioeconomic distributions of canine samples obtained through the passive vs. active rabies surveillance systems in Arequipa, 2021-2022.** The plots in **panel a** show the absolute counts by socioeconomic status (SES) level for canine samples obtained in 2021 and 2022. **Panel b** shows the proportional distributions of samples by SES level.

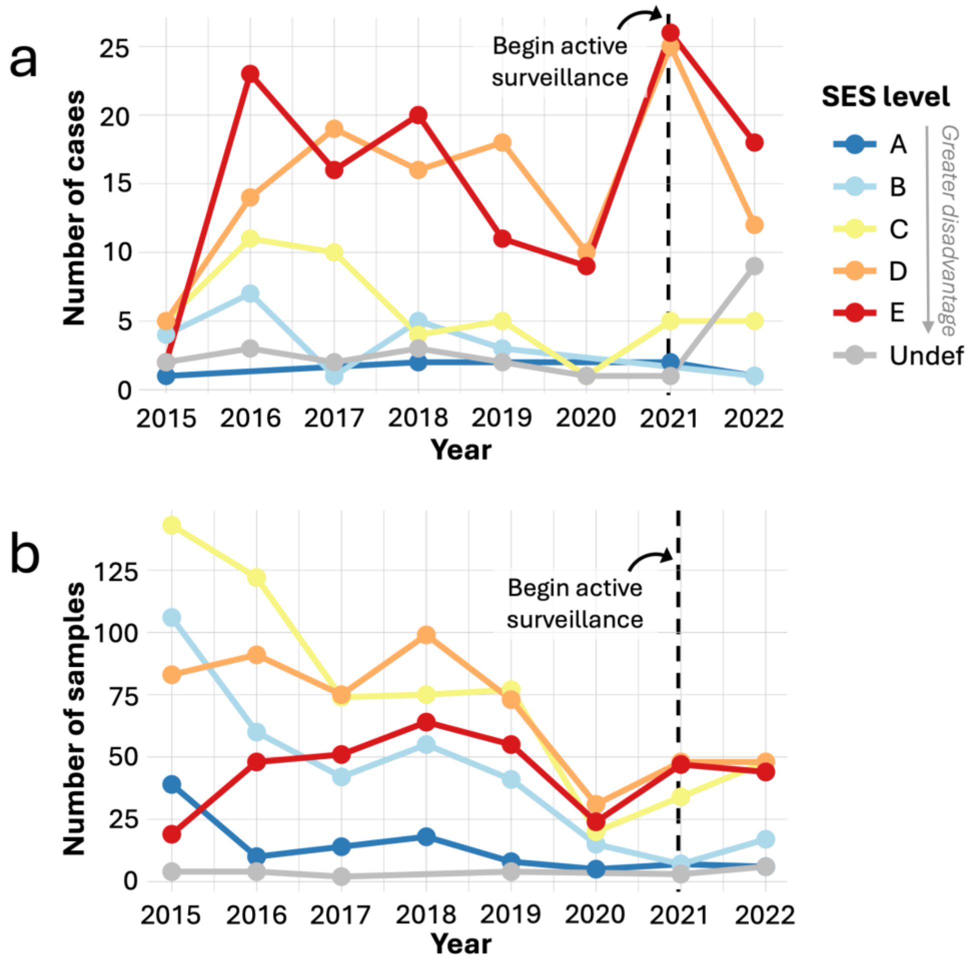

**Figure S3. Temporal trends in the number of confirmed cases (a) and submitted samples (b) by socioeconomic status for canine samples obtained through passive surveillance only.** Compared to Figure 2 in the manuscript, these plots exclude the 153 canine samples and five confirmed cases that were obtained via active surveillance in 2021-2022.

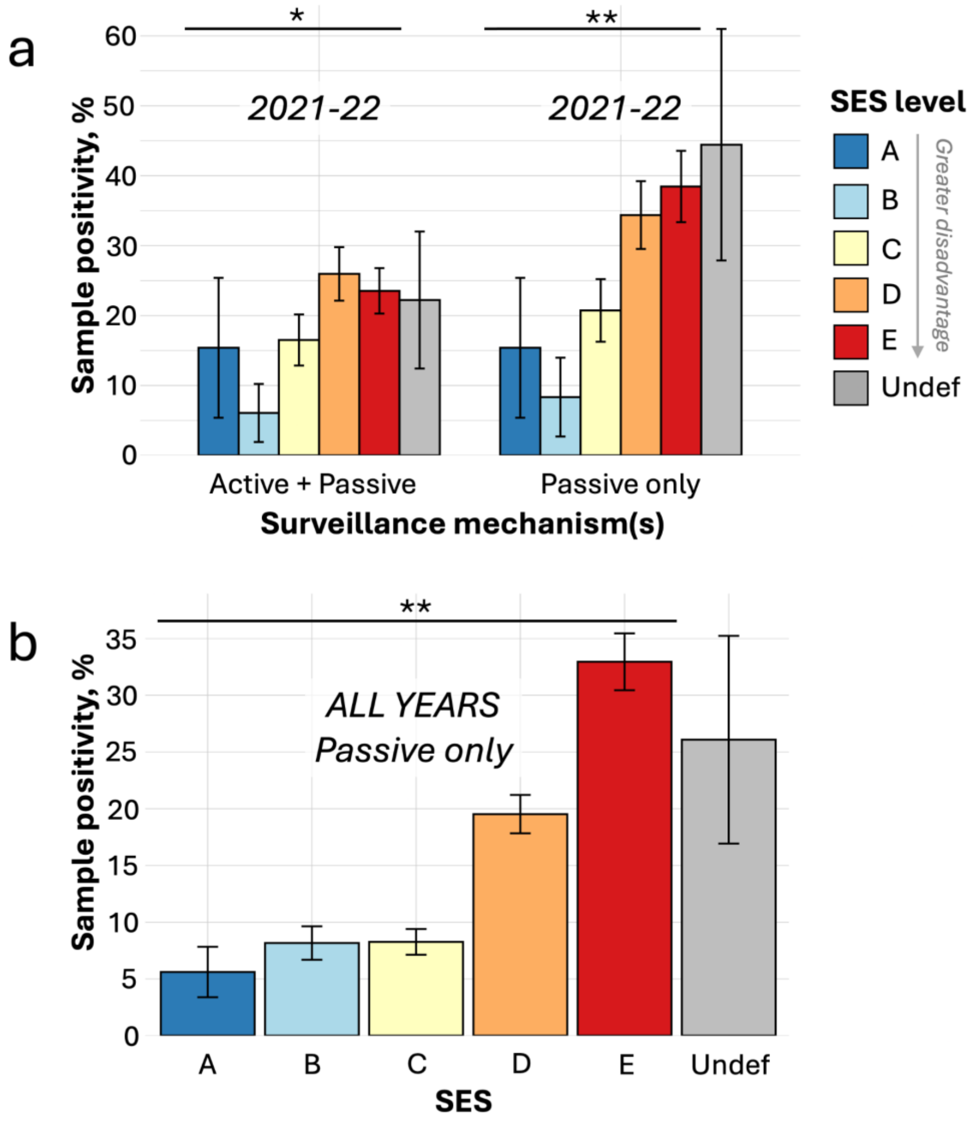

**Figure S4. Comparison of sample positivity vs. neighborhood disadvantage results when samples obtained via active surveillance are excluded. Panel a** shows sample positivity across different socioeconomic status (SES) levels for 2021-22 when active and passive surveillance are included (main results) vs. when active surveillance samples are excluded. **Panel b** shows sample positivity across all years for samples obtained via passive surveillance only. Bars are plotted with their margins of error and shaded according to the SES level associated with the sample of origin. The results of Cochran-Armitage trend tests for a positive association between sample positivity and neighborhood disadvantage (SES A → E) are indicated by asterisks: ** denotes $p$ <0.001 and * denotes $p$ < 0.05.
